# Tumor and pan-tumor diversity and heterogeneity of cancer tissue microbiomes: a medical ecology analysis across 32 cancer types

**DOI:** 10.1101/2024.03.17.24304446

**Authors:** Zhanshan (Sam) Ma

## Abstract

Diversity and heterogeneity are hallmarks of any ecosystems including cancer ecosystems. Tumor heterogeneities have been a hot spot in cancer research because of their critical roles in promoting clonal evolution and metastasis of cancer cells and in influencing cancer progression and therapy efficacy. Cancer tissue microbiome as part of tumor microenvironment can influence tumor heterogeneities both directly and indirectly through their “intimate” intracellular and intercellular interactions with human cells including tumor, immune and normal cells. From an ecological perspective, the relationship between tumor microenvironment and tumor heterogeneity is not unlike that between habitat heterogeneity and community heterogeneity. That is, their heterogeneities should be interwoven with each other, and then the net effects of the microbiomes on cancer development, malignant progression, and therapy responses can be either promotive or suppressive depending on the so-termed immune-oncology-microbiome axis or trio. The objective of this study is to assess and interpret the heterogeneity and often conflated diversity of tumor microbiomes. Our findings, from reanalyzing a big microbiome dataset originally distilled from the TCGA (The Cancer Genome Atlas) database by Poore *et al*. (2020, Nature) including 16555 tumor microbiome samples from the primary tumor (PT), solid tissue normal (SN) and blood derived normal (B) of 32 cancer types, include: (*i*) The tumor microbiome heterogeneity (diversity) cancer relationship HCR (DCR), *i.e.,* the heterogeneity (diversity) differences between PT *vs.* SN (B) are only significantly in approximately 10%-40% depending on the cancer types. (*ii*) The pan-tumor HCR (DCR), *i.e.,* microbiome heterogeneity/diversity differences of same tissue type (*e.g*., PT) across cancer types (e.g., lung vs. breast cancers) are approximately twice the range of previous tumor-HCR (DCR) (*i.e*., 30%-80% for pan-tumor *vs*. 10%-40% of tumor scale). In both tumor and pan-tumor scales, the heterogeneity differences ranges are wider than the diversity ranges. (*iii*) The NSR values range between 0.4 and 0.8 and in 75% cases NSR>0.5, suggesting that tumor selection plays a dominant role than stochastic drifts in shaping microbiome diversity/heterogeneity patterns. Furthermore, the NSR values are significantly different between PT and NT (B) in 50%-100% (mostly 70%-80%) cases across 32 cancer types, further confirming that it should be the tumor growth that is largely responsible for the dominance of selection forces. Finally, we postulate that the HCR (DCR) should be dynamic with tumor types, progression, microbial taxa, host genomics and physiology, therapy and diets.

## 1. Introduction

### 1.1 Heterogenous microbiomes as an emerging cancer hallmark or enabling factors for acquiring hallmarks

Douglas Hanahan (2022) presented a comprehensive review on the state-of-the-art research in cancer hallmarks, which he had co-proposed more than a decade ago and can be defined as a collection of functional abilities gained by human cells as they transform from normal functioning to cancerous growth, particularly traits that are important for their potential to form malignant tumors. In short, cancer hallmarks can be considered as biological capabilities that support tumor development and progression. The eight hallmarks currently encompassing include sustaining growth signals, evading growth suppression, resisting cell death, enabling unlimited replication, inducing or accessing blood vessels, activating invasion and metastasis, reprogramming metabolism, avoiding immune destruction, altering cellular metabolism and avoiding immune destruction. In the updated version, Hanahan (2022), besides arguing that phenotypic flexibility and disrupted development may constitute a separate hallmark ability, he also suggested that non-mutational epigenetic reprogramming and polymorphic (heterogenous) microbiomes can be distinctive enabling characteristics that facilitate the acquisition of hallmark capabilities. His review also examined sources of heterogeneity in cancers and the role of the microbiome. The microbiome can impact cancer risk and progression through mechanisms like DNA damage, proliferation signaling, and immune modulation. Polymorphic variations in microbiomes, *i.e.,* heterogenous microbiomes, between individuals can profoundly impact cancer phenotypes by differentially affecting hallmark capabilities. The microbiome is also a source of heterogeneity between patients.

Lythgoe et al. (2022) further discussed the polymorphic microbes as an emerging hallmark of cancer, while Hanahan (2022) considered microbiomes as enabling factors for acquiring hallmarks. Microbes may be directly carcinogenic (but few species are directly involved), modulate immune responses to promote or suppress cancer, and impact response to therapy. Studies show that the microbiome plays an important part in tumor formation, cancer cell differentiation, and how fast cancer progresses. Additionally, the microbiome interacts with and impacts other established characteristics of cancer, like tumor inflammation, evading immune system destruction, genetic instability, and resistance to cancer treatments. The strongest evidence for the integrated role of the microbiome comes from studying microbes in the gastrointestinal tract, which is the most thoroughly characterized area. However, lately there is growing recognition that polymorphic microbes in other tissues and organs, including other mucous membranes exposed to the external environment (such as skin, genitourinary tract, lung) and those living inside tumors, also play a role (Lythgoe et al. 2022).

### 1.2 Tumor microbiome and pan-tumor microbiome heterogeneities are aspects of tumor heterogeneity from a ‘host’ (tumor) perspective and are microbial community, metacommunity and landscape heterogeneities from a microbial perspective

Intratumor heterogeneity (ITH) refers to cellular diversity within a single tumor, with different cells showing variations in their genetic, epigenetic, and phenotypic profiles; it can influence the tumor’s growth, response to treatment, and potential for metastasis (Marusyk et al 2020 & Peer et al. 2021). According to is Marusyk *et al* (2020) & Peer *et al*. (2021), the ITH arises from genetic, epigenetic, and microenvironmental factors that together generate phenotypic diversity within tumors. Genetic heterogeneity stems from mutations occurring during cell division as well as chromosomal instability. Epigenetic heterogeneity is shaped by differentiation states and environmental cues. Microenvironmental heterogeneity results from disorganization of tumor architecture. ITH increases the odds that some cells will survive therapy and enables ongoing diversification during treatment, leading to resistance. Resistance can involve pre-existing genetic variants, epigenetically plastic persisting cells that later acquire mutations, or microenvironmentally mediated effects. Quantifying ITH could improve treatment strategies by combining drugs, reducing ITH through epigenetic therapies, or using adaptive treatment schedules (Marusyk *et al* 2020 & Peer *et al*. 2021). In contrast, intertumor heterogeneity refers to the differences between tumors in different patients or even different tumors within the same patient. Studies show that tumors vary significantly between patients and can be grouped into distinct categories. The molecular categories represent the genetic mutations, chromosomal changes, epigenetic alterations, and gene expression patterns present in the tumor. These molecular characteristics are more diverse than what can be identified through standard tissue examination. Grouping tumors based on their molecular profiles, which are often closely linked to different outcomes, allows for more precise prognostication compared to traditional classification methods (Marusyk et al 2020 & Peer et al. 2021).

Kashyap et al (2022) argued that understanding heterogeneity across different scales—from single cells to whole tumors and patients—is important for improving diagnosis, prognosis and therapy selection. Recent technological advances now enable quantification of heterogeneity through various data acquisition methods at the molecular, cellular, tissue and organ levels. At the molecular level, techniques like single-cell sequencing, spatial transcriptomics and mass cytometry profile individual cells, while bulk methods like genomic sequencing provide average measurements obscuring rare subpopulations. Computational metrics are then applied to quantify heterogeneity using statistics, information theory or spatial analyses. Radiomics similarly extracts features from medical images to measure heterogeneity. The challenges remain regarding data integration, standardization and clinical validation of metrics. In particular, incorporating multi-omic, multi-scale data through computational frameworks should be able to facilitate the implementation with AI technology for precision oncology. Gough et al. (2017) discusses the need for standardized metrics to quantify heterogeneity in the context of tumor heterogeneity. They propose adopting three metrics: quadratic entropy to measure diversity, the Kolmogorov-Smirnov test to assess normality, and percentage of outliers. Together these form the Pittsburgh Heterogeneity Indices. Other discussed metrics include Shannon entropy, Gaussian mixture models, mutual information, and principal components analysis. They also discuss micro-heterogeneity vs. macro-heterogeneity, spatial heterogeneity vs. temporal heterogeneity, and functional phenotypic heterogeneity (Gough *et al*. 2017, Kashyap *et al* 2022).

Fig 1 illustrated the concepts of tumor and pan-tumor heterogeneity. The term of pan-tumor heterogeneity, which refers to the heterogeneity between different cancer types (such as lung cancer and breast cancer) is a new addition in this paper, which could be considered as part of inter-tumor heterogeneity (e.g., Marusyk *et al* 2022, Peer *et al*. 2021, Kashyap *et al*. 2022). We suggest using the term pan-heterogeneity for inter-cancer type studies (*e.g*., lung *vs*. breast tumors) exclusively. In other words, the primary tumor types are from different cancer types. Some researchers consider tumor microbiome as part of TME (tumor microbiome environment). In this article, we designate tumor microbiome heterogeneity as a separate aspect of tumor heterogeneity. The advantage from having an independent domain for tumor microbiome heterogeneity (TMH) is two-fold. First, the approaches suitable for investigating TMH can be different from the ones used for studying of tumor heterogeneity, given that two domains have different heterogenous objects to investigate, that is, cells *vs*. microbes (archaea, bacteria & viruses). Second, the corresponding approaches in both domains can be different for both historical and technological reasons. The independent treatment allows us to maximally leverage the state-of-the-art research in measuring and interpreting microbiome heterogeneity across microbial communities, metacommunities and landscapes. For example, in the scheme of Fig 1 (Panel A-C), tumor can be treated as “hosts” of tumor microbiomes, which is not unlike the relationship between animal gut microbiomes and their hosts (*e.g.,* Ma 2021). In the meantime, we recognize that tumor tissues have their own characteristics that are different from organism (host) as a whole. Indeed, they can be considered as part of TME, similar to the relationship between habitats and ecological communities of organisms. Putting together, we realize that the biggest challenge in studying tumor microbiome heterogeneity (TMH) is not about how to measure them; instead, the bigger challenge is to establish their relationship with different stages or types of tumors *per se.* For this reason, we conduct extensive comparative analyses across both scales of microbial ecological structures (community, metacommunity, landscape) and of tumors (intra-tumor, inter-tumor, and pan-tumor).

**Fig 1.**
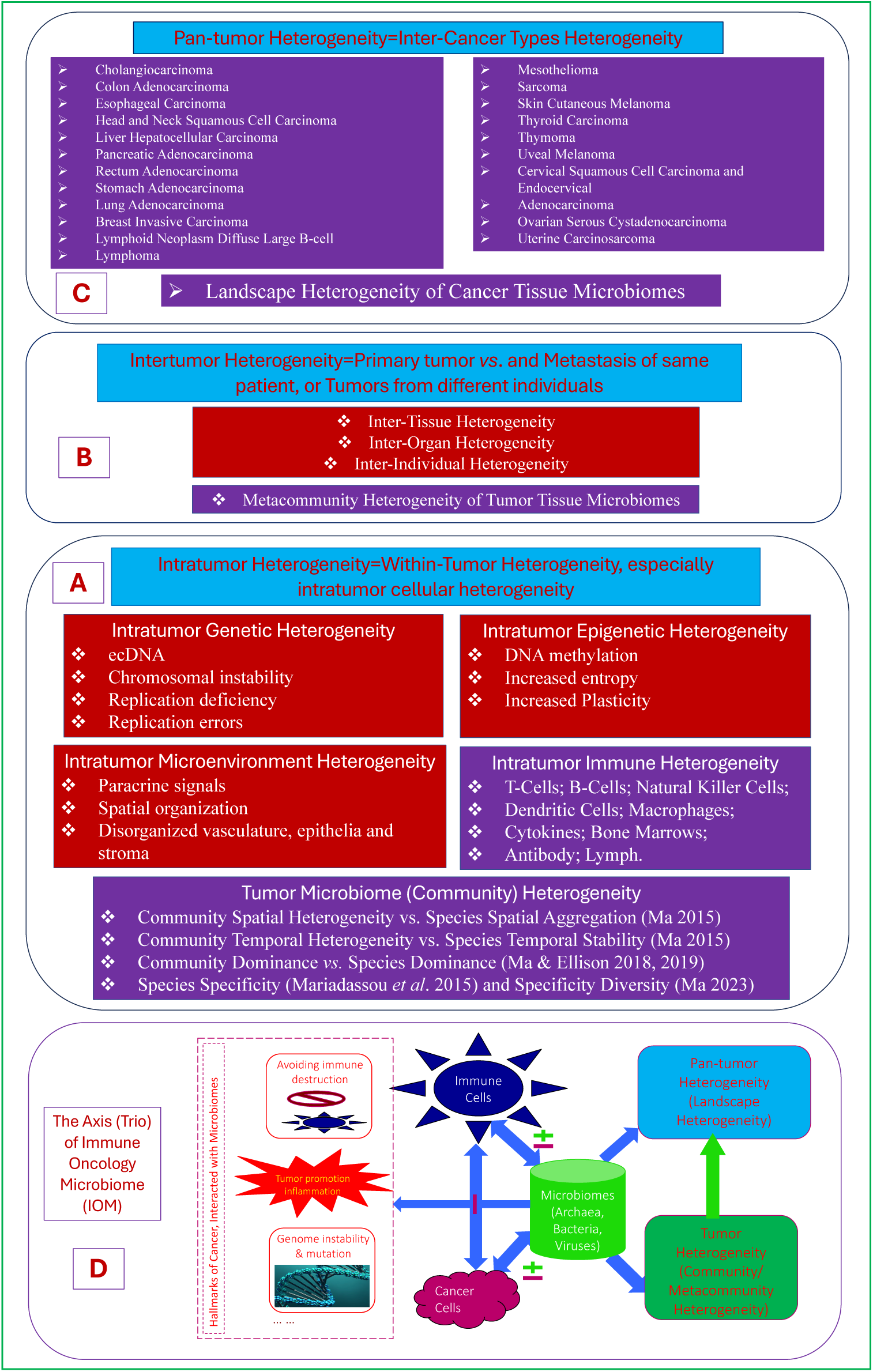
The relationships between tumor microbiome heterogeneity (TMH) and tumor heterogeneity (TH), as well as the IOM (immune-oncology-microbiome) trio. The pink blocks (*e.g*., tumor microbiome heterogeneity, pantumor heterogeneity are new additions to the tumor heterogeneity concept defined by Marusyk *et al*. 2020). The middle blocks (B & C) are inspired by Figure (1) of Marusyk *et al*. (2020). The IOM trio (axis) follows Sepich-Poore *et al*. (2021).

### 1.3 Measuring tumor tissue microbiome heterogeneities across microbial community, metacommunity and landscape

Understanding the heterogeneity of human microbiomes (microbiota) was designated as a mission of the human microbiome project (HMP Consortium 2008, iHMP 2012). Nevertheless, in our opinion, advances in measuring microbiome heterogeneity have been slow, especially in the investigation of the relationship between heterogeneity and diseases (Ma *et al*. 2019, Ma 2020). The biggest challenge in our opinion is the lack of systematic approaches for measuring microbiome heterogeneity. One issue has been the lack of clear conceptual distinction between heterogeneity and diversity, both of which are frequently conflated with each other. A consequence is that much of the claimed heterogeneity studies have been studies on diversity in practice. Diversity, especially biodiversity is essentially the number of species or species equivalents in a community, which can be considered as ‘discrete’ numbers as measured by the Hill numbers (Hill 1973, Chao *et al*. 2014). Diversity, in its essence, ignores interactions between species, while heterogeneity should address interactions, at least, implicitly or indirectly. Heterogeneity should capture the consequence of interactions at the minimum or measure the interactions directly or indirectly. As summarized by Aaron Ellison and Ayelet Shavit in a motto: “a zoo is diverse whereas an ecosystem is heterogeneous,” which encapsulates the distinction between diversity and heterogeneity (Shavit & Ellison 2021). Measuring diversity is not unlike how a zookeeper enumerates her different animals. Her job is relatively easy since she does not need to worry possible interactions between artificially separated animals in a zoo. In contrast, measuring heterogeneity is complicated by interactions such as predator-prey relationships and spatial variation (such as habitat heterogeneity) in ecosystem (Shavit & Ellison 2021). Besides the distinction in dealing with heterogeneity, other slightly less distinctive characteristics include: heterogeneity always involves groups of entities and is often, or by default, associated with space. In the meantime, the conflation between heterogeneity and diversity is also a reality in existing literature, partly because researchers often borrow metrics from diversity research to measure heterogeneity. For example, entropy, which is a standard metric for measuring diversity, is sometimes borrowed to measure heterogeneity (*see* review by Kashyap et al 2022), which is not without merits since unevenness (evenness) can indeed be a proxy of heterogeneity, although often a weak proxy. This is because both unevenness and heterogeneity are influenced by interactions.

The objective of this study is to provide a comprehensive analysis on the heterogeneity and diversity of tumor microbiomes across microbial community, metacommunity and landscape scales. The analysis can be mapped to intra-tumor, inter-tumor and pan-tumor heterogeneities from a tumor (or ‘host’) perspective. The significance of the duo perspective is that it provides a lens to observe the implications of tumor tissue microbiomes to tumor heterogeneity. The latter is known to be of critical importance in understanding cancer progression and therapy response, and more generally in understanding the immunity-oncology-microbiome (IOM) axis or perhaps more accurate the IOM trio (Fig 1, panel D), which highlights the mostly bidirectional triangular relationships between tumor, microbiome and immune cells in cancer ecosystems.

## 2. Material and Methods

Brief descriptions on the cancer tissue microbiome datasets and the analysis design are summarized in the following Box 1. Regarding the metrics and models applied for performing the analyses, Hill numbers (Hill 1973, Chao *et al*. 2014) are well recognized as the most appropriate metrics for measuring alpha-diversity as convincing reasoned in Chao *et al*. (2014), Taylor’s power law (TPL) (Taylor 1961, Taylor 1984) and its extensions (TPLE) for community-level heterogeneity analysis (Ma 2015, Ma & Taylor 2020) are suitable for heterogeneity analysis. We further extended TPL/TPLE onto complex networks to directly address the species interactions, which are the core elements of heterogeneity and also distinguish heterogeneity from diversity, as argued in the introduction section. Ohlmann et al. (2019) network diversity in Hill numbers consist of three metrics: (1) Network diversity of relative abundance (NDRA), (2) Network diversity of link probability (NDLP), (3) Network diversity of link abundance (NDLA). What are measured by the NDLP and NDLA are essentially heterogeneity rather than diversity. Ning *et al*. (2019) normalized stochasticity ratio (NSR) framework is applied to evaluate community and metacommunity stochasticity, which can offer insights on the mechanisms of heterogeneity. Virtually all of the methods except for Taylor’s power law on network (TPLoN) are commonly used metrics/models in medical ecology and their implementation software programs can be found in existing publications such as Chao et al. (2014), Ma (2015), Ning et al. (2019), Ma & Taylor (2020). Detailed computational algorithms/procedures for implementing TPLoN are provided in Box 1.

Before proceeding to present our results, there is a minor, but potentially confusing, terminology issue that should be resolved. As illustrated in Fig 1, in existing literature of tumor heterogeneity, the term “intra-tumor heterogeneity” is used to represent the within-tumor difference, *i.e*., the heterogeneity within a single tumor or single type of tumors, collected from a single individual (patient). In contrast, the term “inter-tumor heterogeneity” is used to represent the between-tumor differences, *i.e*., the heterogeneity between different tumor types (*e.g.,* primary tumor *vs.* metastasis) of same individual, or between tumors from different individuals. If we try to, precisely and directly, align the tumor tissue microbiome heterogeneity with tumor heterogeneity, we may introduce a set of potentially confusing terminology. For example, comparing PT (primary tumor) and SN (solid tissue normal) should be termed “inter-tumor microbiome heterogeneity” because both tissues are certainly of different types. However, we cannot define a counterpart “intratumor microbiome heterogeneity” because another tissue microbiome sample would be from a different individual, which would be “intertumor microbiome heterogeneity” because they are from different individuals. For this reason, in this article, we simply use the term “tumor microbiome heterogeneity” without using “intra” or “inter” prefix for any heterogeneities from comparing the microbiomes collected from the same tumor type (disease) such as lung cancer tissues. Instead, we use the term “pan-tumor microbiome heterogeneity” (or “pan-cancer microbiome heterogeneity”) to refer to the heterogeneities from comparing microbiome samples collected from different cancer types (*e.g*., lung cancer *vs.* breast cancer tissue microbiomes). Our choice is not perfect since intra-tumor microbiome heterogeneity is certainly definable if multiple tissue (*e.g*., PT) samples from same person individual patient, which is not the case in this study. Still our laissez-faire approach is adequate for this study *per se*, does not contradict with existing studies, and is adaptable for future expansions.

In summary, when we compare PT (primary tumor) with SN (solid tissue normal) or B (blood derived normal) of same cancer type (*e.g*., lung cancer only), either from single patient or cohort of patients of the same cancer, we use the term “tumor microbiome heterogeneity;” when we compare metrics and/or model parameters calculated from samples of different cancer diseases (e.g., lung cancer *vs*. breast cancer), we use the term “pan-tumor microbiome heterogeneity.” In pan-tumor comparisons, we only make ‘counterpart’ comparison (*e.g*., PT of lung cancer with PT of breast cancer), without comparing different types of samples (e.g., PT of lung cancer with SN of breast cancer) for obvious reason.

**Box 1**. Brief descriptions on the cancer-tissue microbiome samples (datasets) and the reanalysis schemes of this study (also *see* Fig 1 for the related concepts).

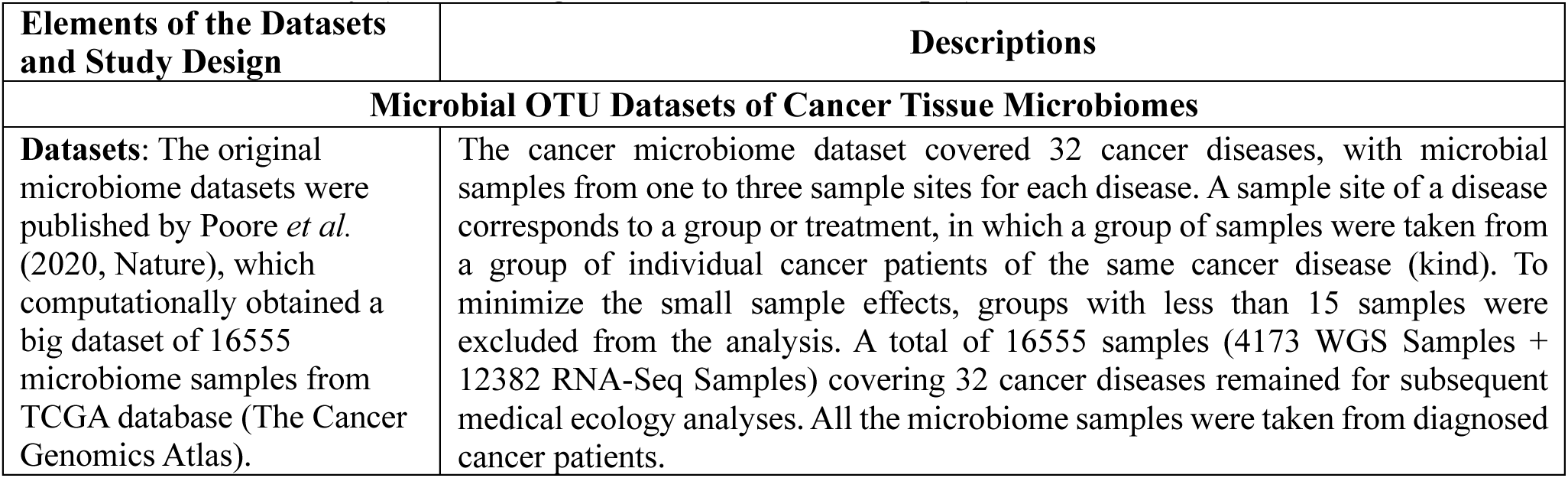
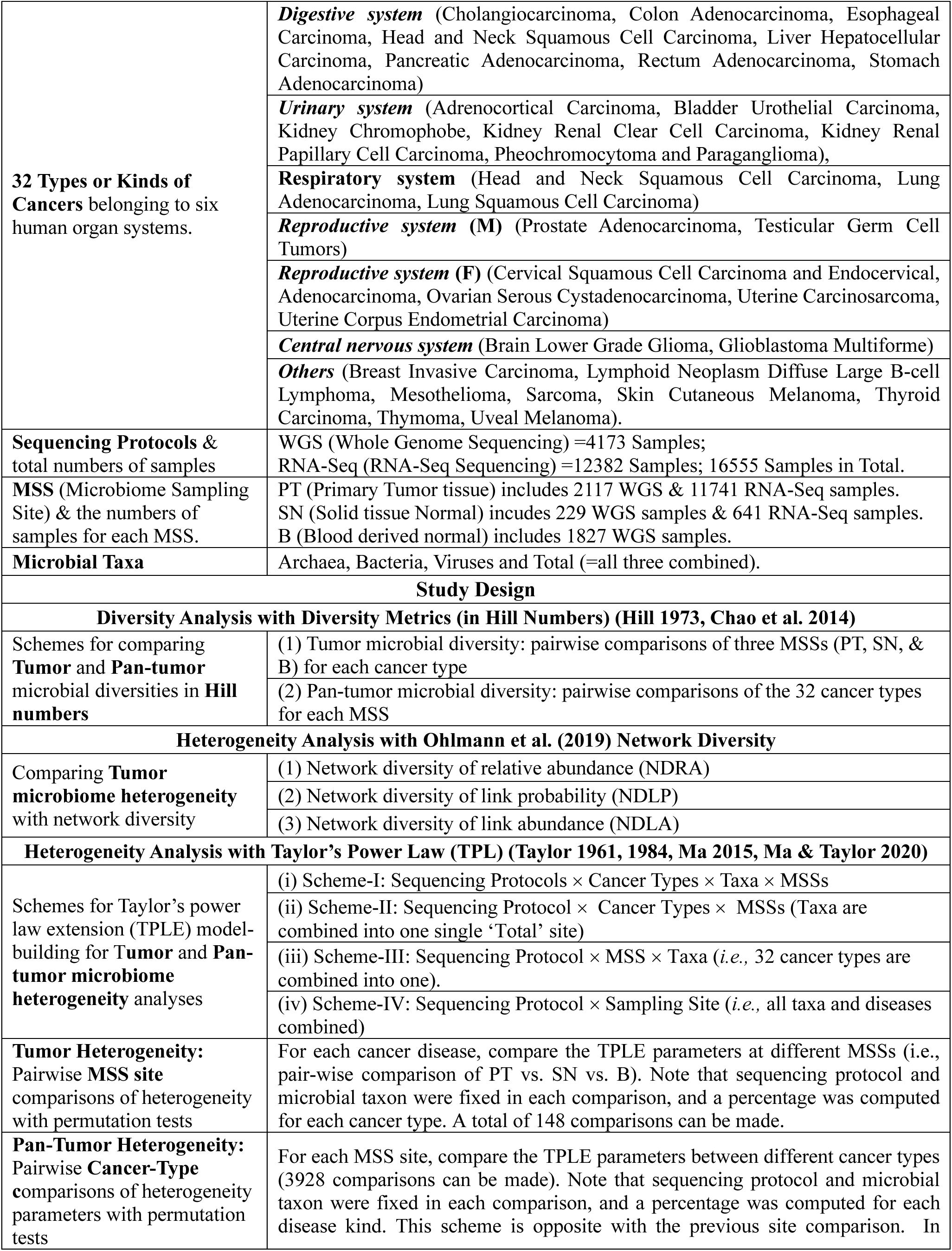
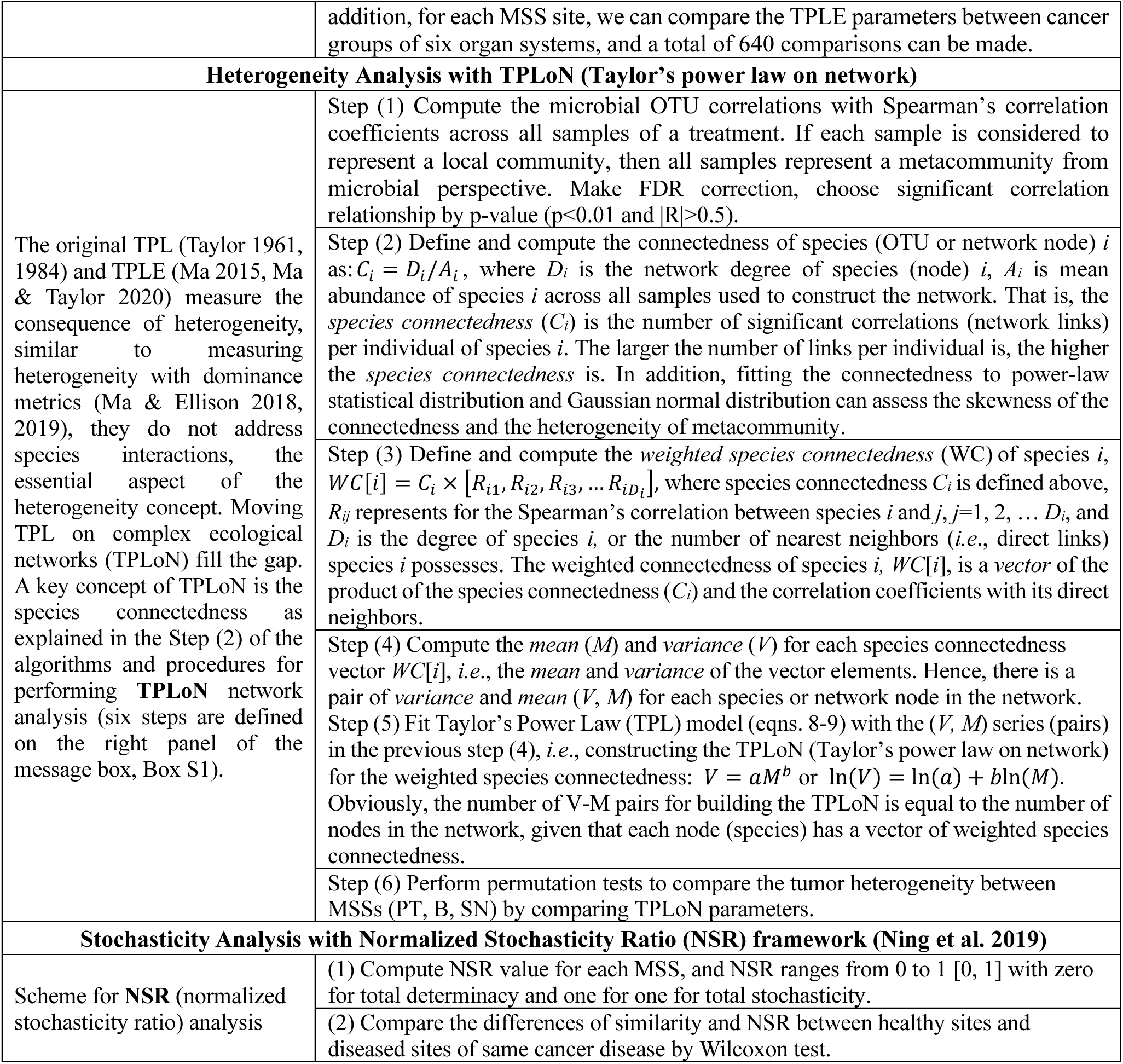

## 3. Results and Discussion

### 3.1 Tumor and pan-tumor microbial diversity analyses with Hill numbers

#### 3.1.1 Comparisons of the tumor microbial diversity between healthy and primary tumor microbiomes across 32 cancer types

Table S3, summarized from Tables S1 for the values of diversity metrics and S2 for the effect sizes from significance tests, exhibits the pair-wise comparisons of the three microbiome sample sites (MSSs) in term of their microbial diversities measured in Hill numbers across 32 cancer types for each microbial taxon (archaea, bacteria, or viruses and their ‘Total’) under two different sequencing protocols (WGS and RNA-Seq) (Fig 1). The three MSSs include solid tissue normal (SN), blood derived normal (B), and primary tumor (PT). The comparisons for diversity (Hill numbers) were made with effect size tests (Cohen’s *d* statistic). The main findings from Table S1-S3 can be summarized as follows:

First, the differences of diversity in Hill numbers between the healthy tissues (SN or B) and primary tumor (PT), *i.e.,* SN *vs*. PT, and B *vs*. PT, mostly range 30%-40% approximately, and the range of differences may be attributed to the differences in taxa (archaea, bacteria and viruses) and sequencing protocols (WGS/RNA-Seq.) (Table S3). It should be noted that the range of 30%-40% stated here is approximate and reflects the variations of majority of comparisons given that the extreme of the range was 0%-72% for a small handful of the comparisons. In the remainder of this article, our quantifications of ranges follow the similar convention to simplify our presentation of the results without losing generality.

The above finding regarding the tumor microbial diversity between healthy tissues (SN and B) and PT across 32 cancer types is consistent with the so-termed 1/3 DDR (diversity-disease relationship) conjecture, which postulates that the DDR is only significant in approximately 1/3 of cases (Ma *et al*. 2019, Ma 2020). Furthermore, the differences in microbial diversity among different microbial taxa (archaea, bacteria, or viruses) or sequencing protocols (WGS vs. RNA-Seq) are approximately 10% on average in most comparisons, and these differences are not sufficiently large to shadow the general 1/3 pattern of the diversity-cancer relationship (DCR) in our opinion. We emphasize that the tumor microbial DCR of 1/3 differences is on average-level of different microbial taxa (including archaea, bacteria, and viruses).

#### 3.1.2. Comparisons of the pan-tumor microbial diversity between different cancer types (32 in total) for each tissue microbiome (B, SN & PT)

Table S6 (summarized from Tables S4 & S5) shows the results (percentages with significant differences) of the effect-size test for microbial diversity (in Hill numbers) based on the pair-wise comparisons of the 32 cancer diseases for each MSS. The differences in microbial diversity between different cancer types (32 in total) or the pan-tumor microbial diversity differences range between 70%-80% approximately, which is almost double size (magnitude) of the differences in tumor diversity as summarized previously in Table S3. Similar to tumor diversity, the diversity differences due to sequencing protocols and taxa are relatively small, and the variations are around 10% in general, which is not sufficiently large to shadow the general patterns of pan-tumor microbial diversity among cancer types.

Table S6 also suggests that the pan-tumor biodiversity differences between different cancer types in PT site is generally more prevalent than those in SN or B site. This should be expected since it simply highlights the effects of tumors on microbial diversity, which can be considered as disturbances to tissue microbiomes and should cause fluctuations of microbiome abundances and distributions. This also explains the observation in Table S6 that the SN usually has the lowest pan-tumor heterogeneity, especially for higher diversity order (*q*=1-3). An interesting observation in Table S6 is that the differences in microbial diversity of blood microbiomes between different cancer types at higher diversity orders (*q*=1-3) appear to be as large as those of PT and to be more prevalent than those of SN. This finding suggests that blood microbiome seems more sensitive than solid normal tissue to tumor disturbances, which may be of important clinical implications. For example, testing blood microbiome changes could be more sensitive than testing solid tissue normal and equally sensitive with testing primary tumor tissues.

#### 3.1.3 Average microbial diversity across cancer types

Table S7 exhibits the averages of microbial alpha-diversity in Hill numbers across all cancer types for each MMS (microbial sampling site, *i.e.,* PT, SN, & B) under each sequencing protocol (WGS or RNA-Seq) and taxon (archaea, bacteria, viruses) (also See Fig 2). The bottom two sections of Table S7 further averages the Hill numbers across taxa and MMSs, respectively.

**Fig 2.**
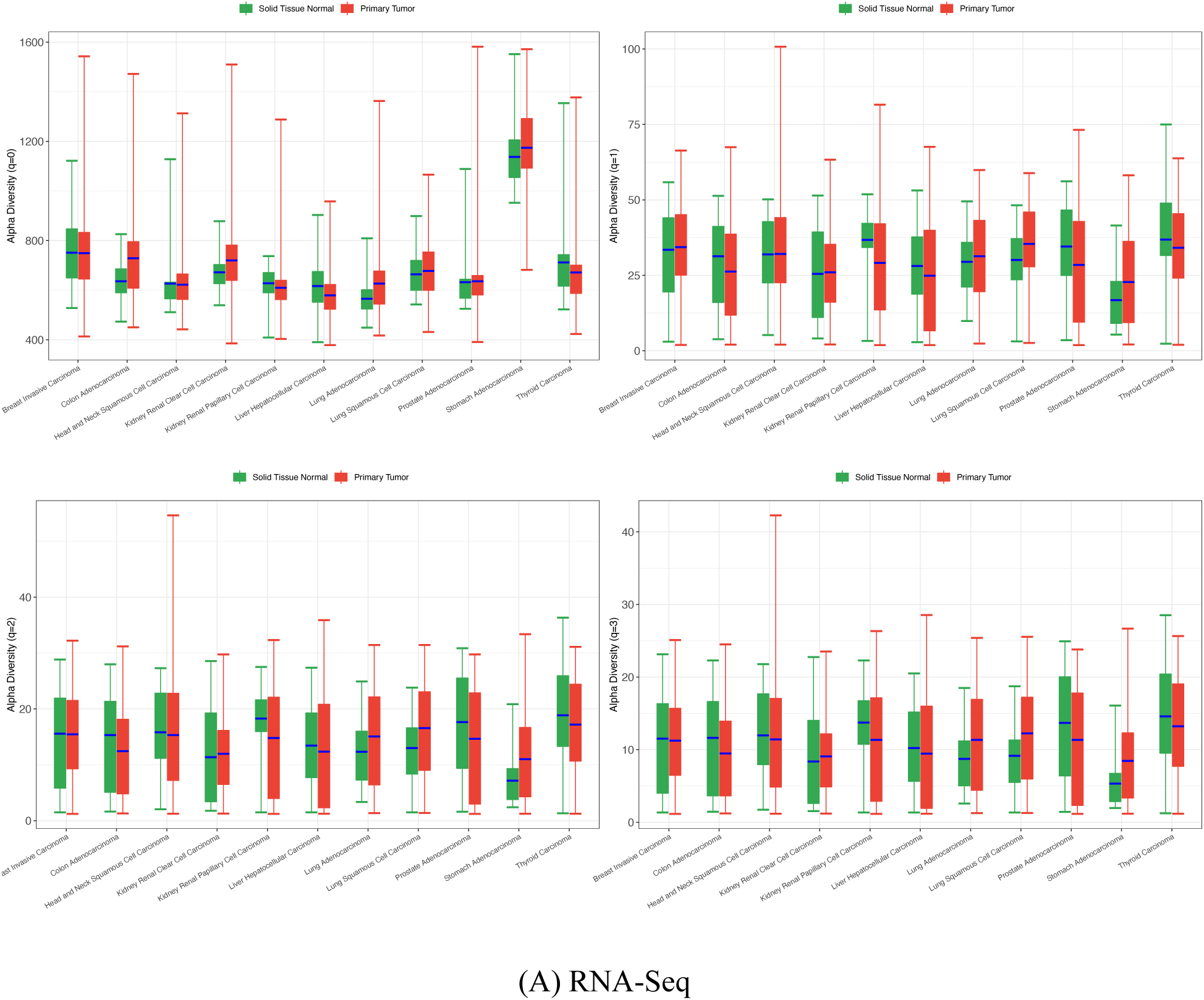
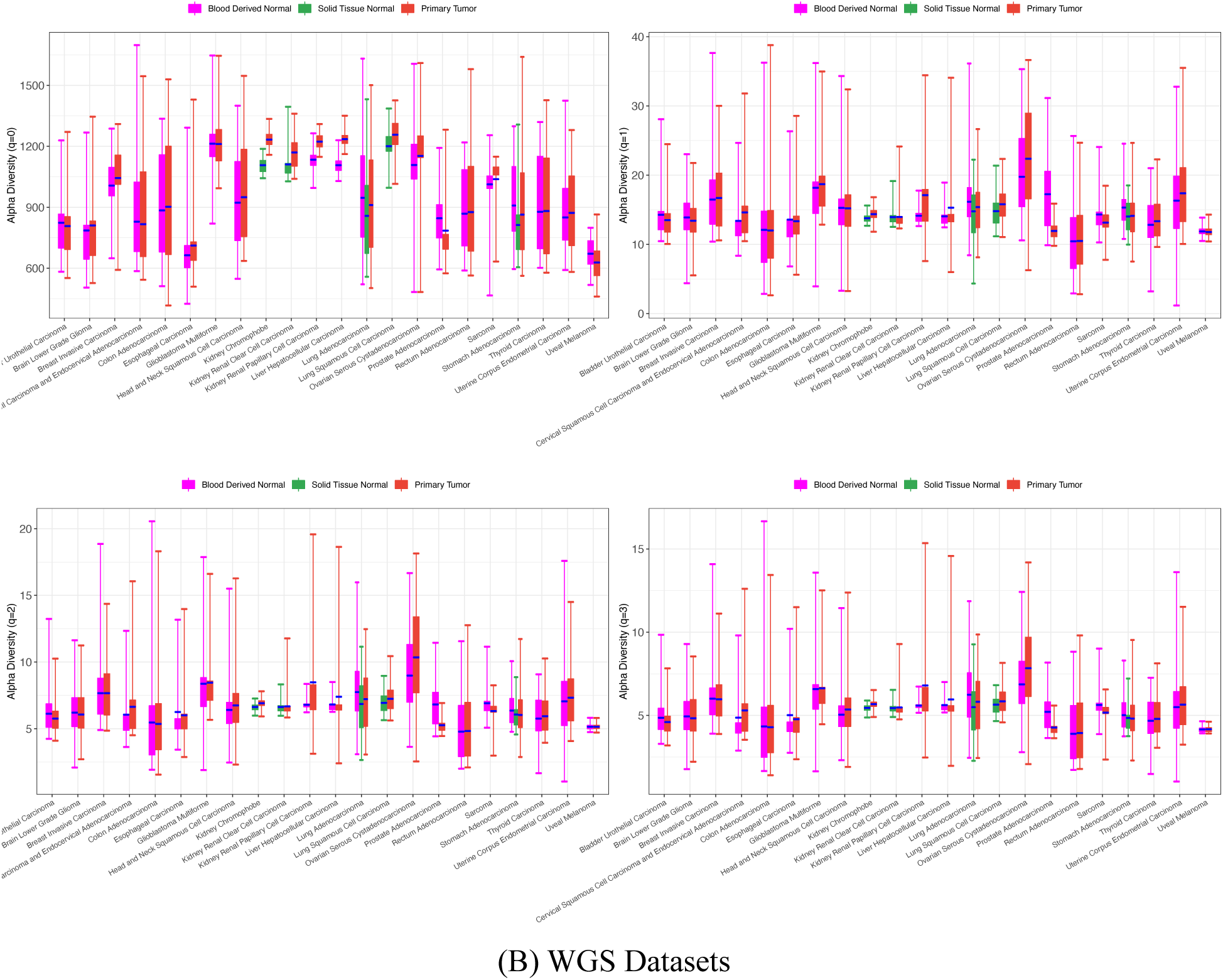
Standard bar charts showing the alpha-diversity in Hill numbers at different diversity orders (*q*=0-3) for the total (of archaea, bacteria, and viruses combined) microbiome of various cancer types: (A) based on RNA-Seq datasets; (B) WGS sequencing datasets.

As to the order of diversity sizes among MSSs, different sequencing datasets produced different patterns. On average, with WGS data, the species richness in SN is higher than in PT, which is higher than in blood, that is, SN>PT>B. However, the diversity at higher orders (*q*=1-4) shows mixed patterns, in most comparisons, PT>SN.

On average, with RNA-Seq data, the species richness (diversity at q=0) in PT is larger than that in SN. However, the diversity at higher orders (*q*=1-4) exhibited the opposite trend, that is, SN>PT. We postulate that two factor may be responsible for this apparent paradox: (i) RNA sequencing data only include the expressed microbial genes; (ii) the positive and negative effects of pan-tumor diversity maybe canceled each other in the averages. For this reason, we suggest taking caution with the averages summarized in Table S7 for inferring general patterns on diversity-cancer relationships. In other words, which MSS site host higher microbial diversity at pan-tumor scale is uncertain, depending on cancer types, possibly also on cancer progression stage, host physiology and genomics, as well as their lifestyle and diet habits.

In summary, the tumor diversity-cancer relationship (DCR) follows the general diversity-disease relationship (DDR) in microbiome associated diseases, which suggested that the relationship is statistically significant in only about 1/3 of cases (Ma *et al*. 2019), *i.e*., 30%-40%. The pan-tumor DCR across 32 cancer types in this study (*i.e.,* 70%-80%) suggest approximately twice the magnitude of tumor DCR. As to the direction (i.e., order of diversity sizes) in DCR relationships, whether cancer enriches or depletes diversity, depends on specific cancer types, at least, and possibly on other factors such as cancer stages, patient physiology and genomics, and their lifestyles and diet habits.

### 3.2 Tumor heterogeneity analysis with network diversity

The so-termed network diversity (Ohlmann *et al*. 2019), which appears to be a misnomer, essentially measures the microbiome heterogeneity, especially the network diversity of link probabilities (NDLP) and network diversity of link abundances (NDLA) that are computed from species interactions (correlations) and the synthesis of correlations and abundances, respectively Fig 3 shows an example of networks used to compute the network diversity.

**Fig 3.**
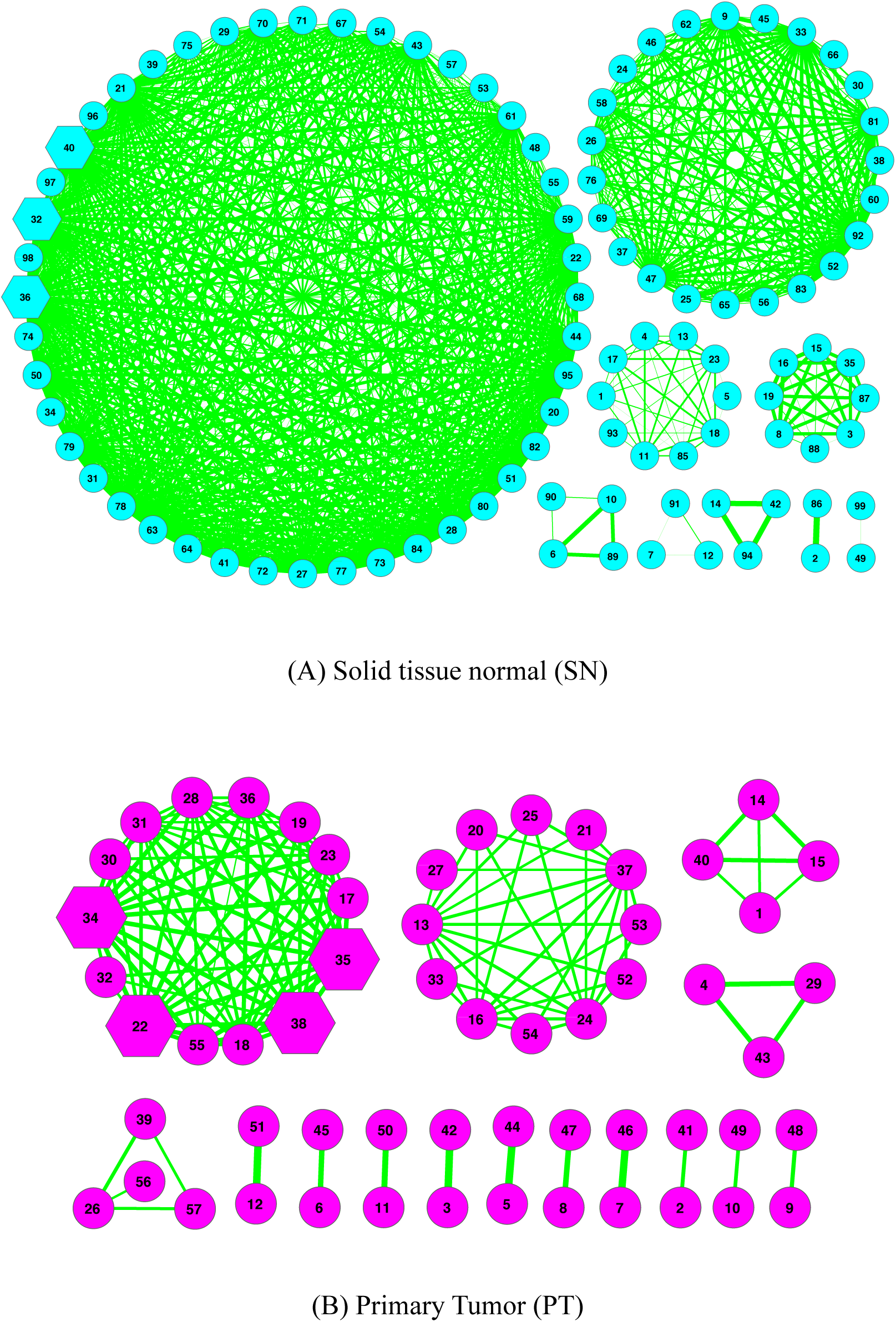
The species cooccurrence networks of tissue microbiome networks of stomach adenocarcinoma patients: (A) Solid tissue normal (SN), (B) Primary tumor (PT).

Table S8 exhibits the network diversity computed for each of the combinations of cancer types (32 in total), MSSs (B, SN, PT), taxa (archaea, bacteria, viruses), and sequencing protocols (WGS vs. RNA-Seq). Table S9 lists the *P*-values of the permutation tests for pair-wisely comparing the network diversities of B, SN and PT microbiomes. Table S10 shows the percentage of comparisons with significant differences in network diversity between MSSs, *i.e.,* the tumor heterogeneity.

To compute network diversity, we first build species cooccurrence networks (*see* Fig 3 for one example) with the microbiome samples of a specific MSS tissue. Fig 3 displays the network graphs of PT and SN of stomach adenocarcinoma. Fig S4 illustrates the network diversity of link abundances (NDLA) of total taxa (archaea, bacteria, and viruses) microbiomes at different diversity order (*q*=0-3) of different cancer types.

With RNA-Seq sequencing data, comparing the differences in network diversity between PT and SN revealed approximately 10%-36% differences, and the magnitude (range of differences) depends on diversity order (*q*), types of network diversity (NDRA, NDLP, NDLA), and taxa. Among the three types of network diversity, the NDRA or relative-abundance diversity is very similar with the traditional diversity, the NDLP or link-probability diversity is the diversity of interactions, which is equivalent with heterogeneity, and NDLA or link-abundance diversity is considered as a synthesis of both NDRA and NDLP. Possibly due to the synthesis or overlap effects between abundance and interactions, the NDLA exhibited highest diversity values among the three types of network diversity. The zero percentage of differences in network diversity occurred with archaea and viruses at diversity order q=3, and none occurred with bacteria at any diversity order.

With WGS sequencing data, comparing the difference in network diversity between B and SN revealed no difference in any diversity orders except for a small number (7 out of 48) of comparisons. Since both B and SN are considered as healthy tissue, the lack of wide differences in microbiome heterogeneity is somewhat expected.

With WGS sequencing data, comparing the difference in network diversity B and PT revealed 10%-36% differences, and the magnitude (range of differences) depends on diversity order (*q*) and the types of network diversity. The differences here are similar with the previous SN vs. PT with RNA-Seq datasets. Also, three types of network diversity did not exhibit a specific order in terms of their sizes (values).

With WGS sequencing data, comparing the differences in network diversity between SN and PT revealed 0%-100% differences, and the huge range of differences varied by diversity order (*q*), types of network diversity and taxa. For the relative-abundance diversity or NDRA, the pattern is similar to previous patterns of B vs. PT in WGS and SN vs. PT in RNA-Seq, which should be expected given the nature of this type of network diversity, which is essentially the traditional diversity in a network setting or node diversity. The difference between NDRA and traditional diversity is that the former excludes those species that are isolated nodes and do not interact with any other species in the network (communities). But such kind of isolated nodes are rare in practice. For the two other network diversity types with WGS data, when *q*=0, the NDLP (or link-probability diversity) and NDLA (link-abundance diversity) ranged from 40%-100%. In contrast, most comparisons (22 out of 24) of SN vs. PT with WGS datasets exhibited insignificant differences when q>0. This is somewhat puzzling.

Compared with previous diversity measures, the differences in tumor heterogeneity between the normal (B or SN) and tumor microbiomes, measured with link-probability diversity (NDLP) and link-abundance diversity (NDLA) appear to fluctuate more dramatically than tumor diversity. Nevertheless, it should be emphasized that heterogeneity and diversity measure different aspects of microbiomes, and direct comparisons of their numbers are not necessarily meaningful.

### 3.3 Tumor and pan-tumor heterogeneity of microbiomes with TPLE

#### 3.3.1 Tumor microbiome heterogeneity with TPLE modeling

Table S11 and Table S12 exhibit the TPLE (Taylor’s power law extensions) model for each of the combinations of cancer types, microbial taxa, and MMSs, for RNA-Seq and WGS respectively. The TPLE models built here are basic cancer type level model, one model for each MMS of each cancer type (actually also for each taxon), and they can be utilized to analyze tumor heterogeneity of microbiome. Table S13 summarized the average parameters of the basic TPLE models in Table S11 and Table S12 across 32 cancer types, and they can be particularly useful for assessing the general parameter ranges of tumor TPLE models, and to some extent, the pan-tumor variability of tumor heterogeneity. We summarize the findings regarding tumor heterogeneity as following points:

The tumor TPLE heterogeneity models fit to the datasets of cancer tissue microbiomes extremely significant (*P*-value<0.001, see Fig 5). All TPLE scaling parameters values (*b*) exceed *1*, suggesting highly heterogenous microbiomes. Majority of community heterogeneity scaling parameter (b≥2) suggest rather heterogenous distribution of microbes at community level (type-I TPLE), while the population-level heterogeneity (type-III TPLE) is relatively small as indicated by (1<b<2) (Tables S11-S13).

**Fig 4.**
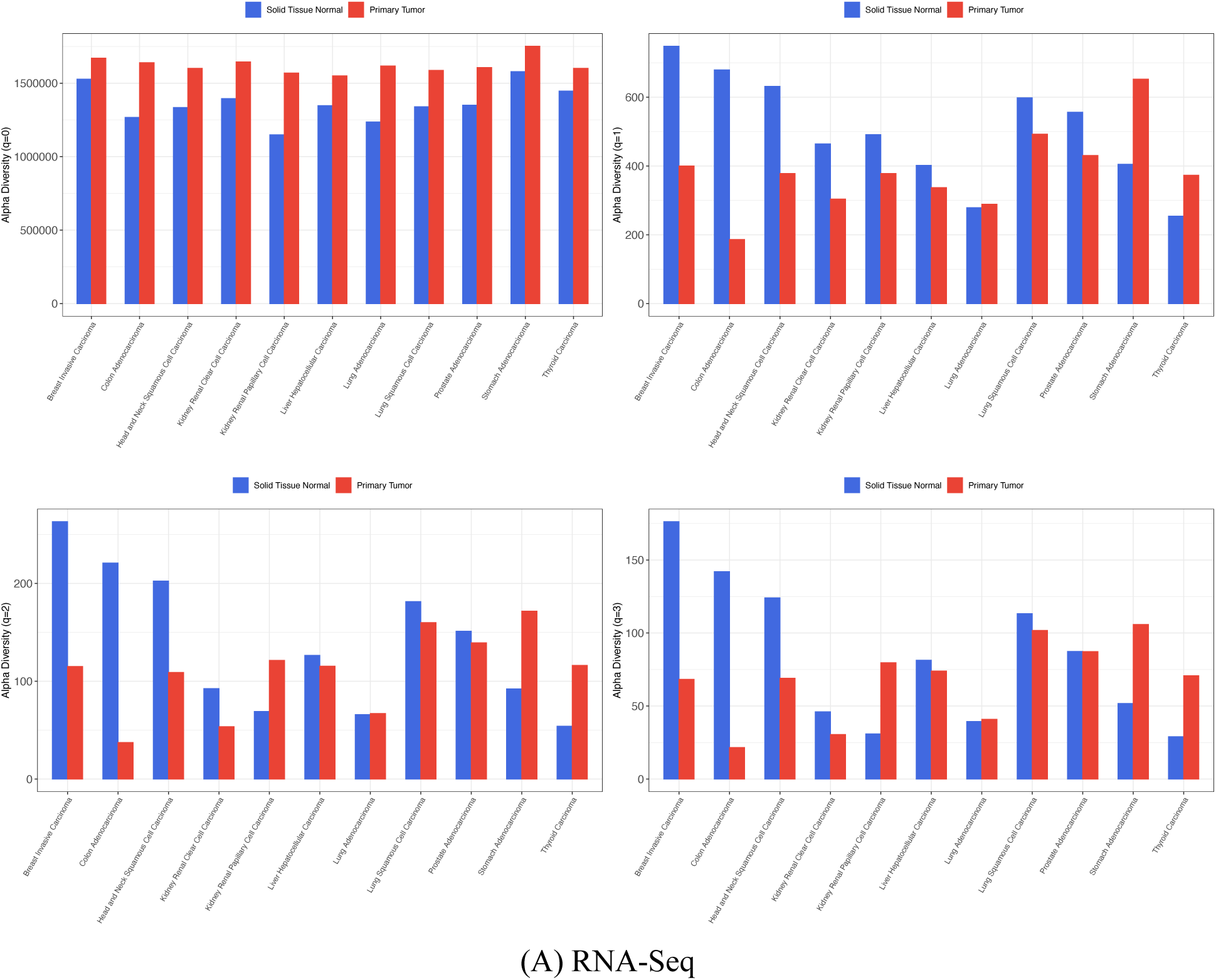
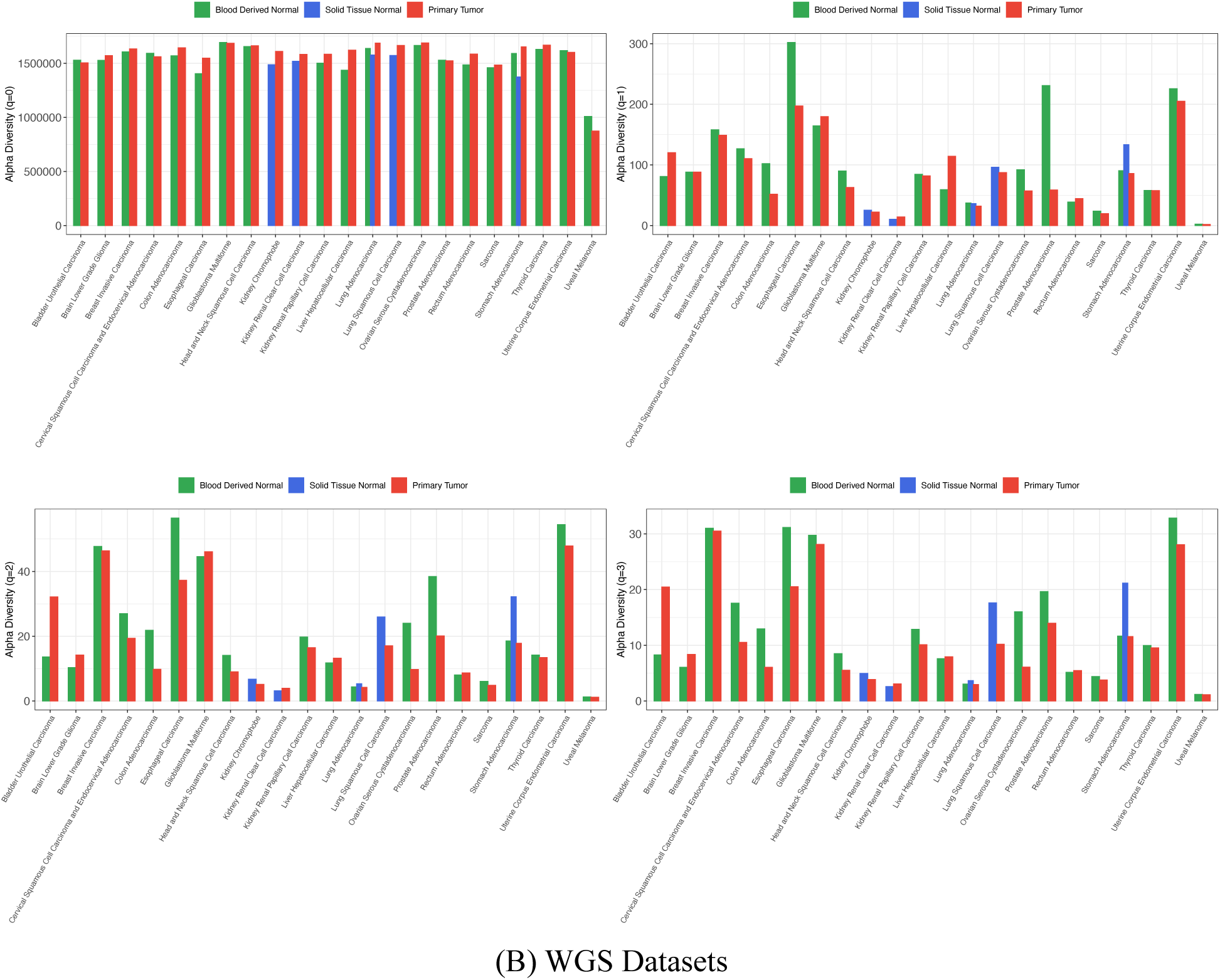
The NDLA (network diversity of link abundance) of total (archaea, bacteria, and viruses) microbiomes at different diversity order (*q*=0-3) of different cancer types: (A) RNA-Seq; (B) WGS.

**Fig 5.**
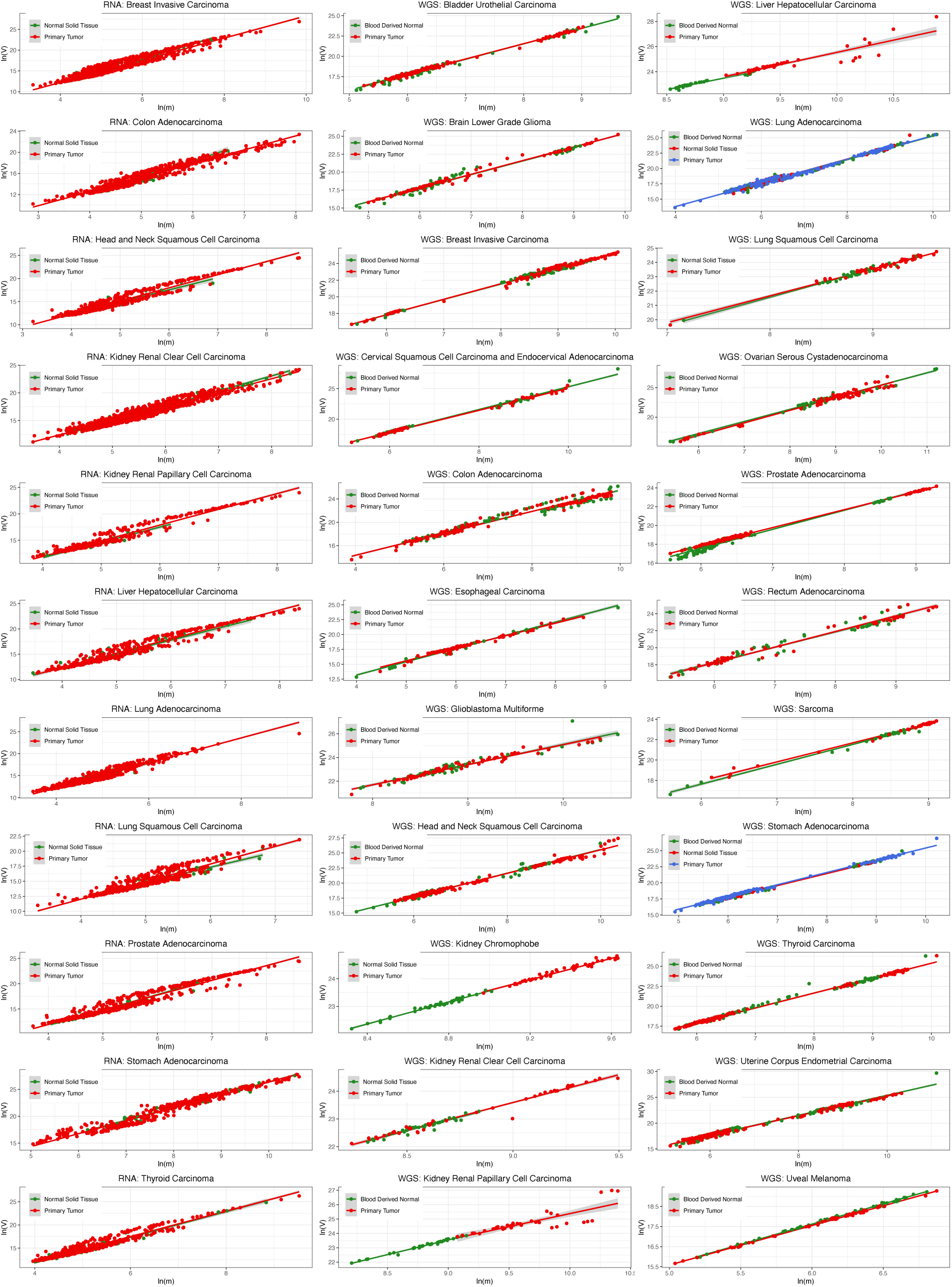
Fitting type-I TPLE (Taylor’s power law) model for measuring community heterogeneity with cancer tissue microbiome datasets

Permutation tests (Table S14) for the pairwise differences of scaling parameter (*b*) between MSSs (B, SN, PT) show that at community scale, the percentage with significant differences ranges between 10%-40%, and at the population scale, the percentage ranges between 10%-50%. The test results in Table S14 do not distinguish between normal (B or SN) and tumor (PT) tissues, and they simply demonstrated the inter-MSS heterogeneity, not necessarily the heterogeneity between the normal (SN or B) and PT microbiomes.

Table S15 further summarized the permutation test results of Table S14 from the perspective of normal *vs*. PT perspective, i.e., B *vs*. PT, SN *vs*. PT, which should be more interesting. But first, the comparison between B and SN revealed no significant differences in heterogeneity (0%) at community scale, and only bacteria showed significant differences of 50% at population scale. This is somewhat expected given that both B and SN are considered as normal tissues. The comparison between B and PT revealed 10%-40% differences approximately at community scale (Type-I TPLE) and 20%-50% at population scale (Type-III TPLE). Furthermore, the comparison between SN and PT revealed approximately 10%-40% differences at community scale and approximately 20%-50% at population scale. That is, the normal (B or SN) vs. PT comparisons revealed similar patterns of differences—10%-40% at community scale and 20%-50% at population scale.

The above-described ranges of differences in tumor heterogeneity can be attributed to sequencing protocol and taxa, which superimpose additional influences on the heterogeneity difference beyond the influence of tumors.

In summary, the tumor heterogeneity differences between PT and normal controls (B or SN) range between 10%-40% at community scale and 20%-50% at population scale. The percentage and range of heterogeneity differences are similar to those of tumor diversity comparisons, but they measure different aspects of tissue microbiomes as argued previously. The finding here is also consistent with the previous finding based on network diversity.

#### 3.3.2 Pan-tumor heterogeneity of microbiome with TPLE modeling

For each MSS, comparing the heterogeneity-TPLE parameters between 32 different cancer types can reveal the pan-tumor heterogeneity. Table S16 exhibit the pan-tumor heterogeneity TPLE models that were built by pooling together the datasets of all cancer types and one model was built for each taxon (archaea, bacteria, viruses, and total of the three taxa) of each MMS (PT, SN, or B) under each sequencing protocol (WGS or RNA-Seq). A total of 48 models were built. All of the pan-tumor heterogeneity TPLE models were fitted to the data extremely significant (*P*-value=0.001). All community scale models are of (*b*-value≥2) and population scale are of 1<b<2, suggesting higher heterogeneity at community scale, although heterogenous at both the scales.

The pan-tumor community heterogeneity TPLE parameter *b* showed approximately 30%-70% differences between different cancer types (Tables S17 & S18). Furthermore, the pan-tumor heterogeneity of primary tumor (PT) is on the large side of the interval (-70%) and that of normal tissue (SN or B) is on the small size of the interval (30-%). The difference of pan-tumor heterogeneity (30%-70%) here is almost twice the level of the previous tumor heterogeneity (10%- 40%). Furthermore, the pan-tumor heterogeneity of PT is larger than that of B or SN, which is again consistent with tumor heterogeneity.

The pan-tumor population-scale heterogeneity (measured in TPLE-b) showed slightly higher difference range (50%-70%) than that of community-level difference range (30%-70%). The differences between PT and normal tissue (B or SN) at population scale is also similar with the previous community-scale pattern, *i.e.,* PT showing larger difference than SN or B (Tables S17 & S18).

Sequencing protocols (WGS or RNA-Seq) and taxa contribute to the range of the pan-tumor heterogeneity parameters, but their influences are limited to the range of the interval, which are relatively small and are not sufficient to shadow the patterns of pan-tumor heterogeneity across cancer types and MSSs.

## 4. Network heterogeneity with TPLoN (Taylor’s power law on network)

The previous heterogeneity analysis with TPLE essentially measures the consequence of species interactions, because what are modelled with TPLE are still species abundances. In this section, by moving TPL onto species cooccurrence networks, we can directly measure the heterogeneity scaling of species interactions, which is measured in species connectedness and computed from species correlation, degree and abundances in TPLoN (see Table S1 for the detailed computational procedures).

Table S19-S20 exhibit the TPLoN parameters for each MSS site of each cancer type based on RNA-Seq and WGS datasets, respectively. Table S21 exhibits the corresponding permutation test for the differences in the TPLoN parameters exhibited in Table S19 (for RNA-Seq) and Table S20 (for WGS) (also see Fig 6). In both sequencing protocols, the network heterogeneity parameter (*b*) exhibits smaller values than in previous non-network or classic TPLE models, where b ≥ 2 in most cancers, while here *b*<2 in most cancers. In general, the parameter (*b*) of TPLoN is smaller than that of non-network TPLE (*P*-value=0.005). This difference suggests that the heterogeneity scaling in network setting (species correlations or interactions are the focus) is slower than in non-network setting (species abundances are the focus). This should be expected given that the species interactions (relationships) are usually much stable than fluctuations of species abundances.

**Fig 6.**
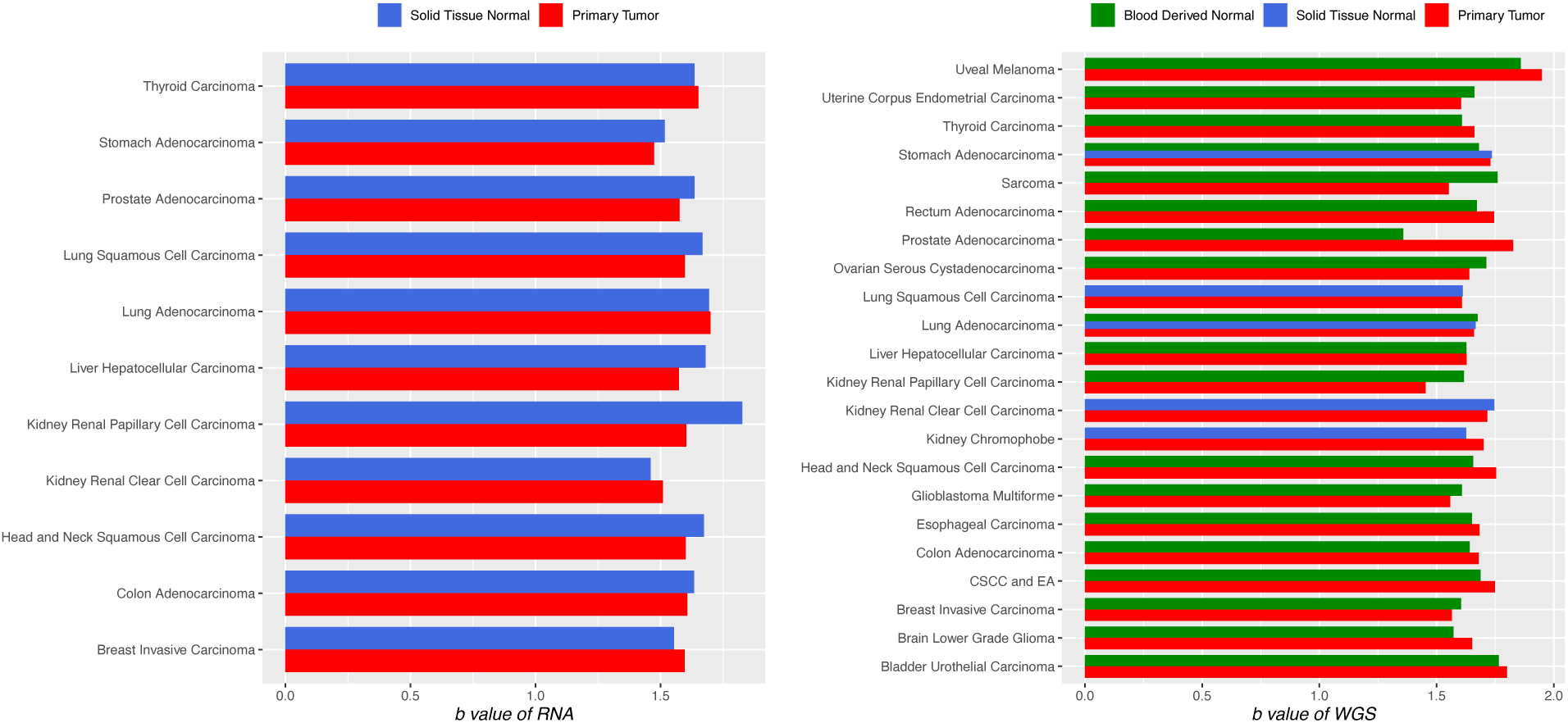
The tumor network heterogeneity scaling parameter (b) of TPLoN (Taylor’s power law on network) built with RNA-Seq (the left) and WGS (the right) datasets.

The permutation tests (Table S21) show that the percentages of comparisons with significant differences between PT and SN or between PT and B range between 9%-12% with WGS sequencing having slightly higher percentage (12%) of differences than RNA-Seq (∼9%). Compared with approximately 10%-40% differences in heterogeneity measured in classic TPLE in previous sub-section, the 9%-12% differences in TPLoN-measured heterogeneity can be considered as ‘pure’ heterogeneity of species interactions, which are, strictly speaking, the heterogeneity of species cooccurrences or distributions across metacommunity. The smaller differences between normal tissues (B or SN) and PT may simply reflect the reality that species interactions should be more stable than species abundances in general.

We further studied the statistical distributions of the *species connectedness* in the TPLoN networks of different MSSs under two different sequencing protocols with power-law statistical distribution and Gaussian normal distribution, respectively. Table S22 and Table S23 exhibit the results from fitting power-law statistical distribution and Gaussian normal distribution. It turned out that in approximately 21%-23% of the cancer types, the species connectedness of TPLoN of PT or blood (B) follows the power-law distribution, and however none of the SN follows the power-law distribution. Furthermore, none of the TPLoN connectedness satisfied with the Gaussian normal distribution. This finding suggests that tumor does influence the species connectedness distribution of heterogeneity network, but the influence is not universal, and its influence is only significant in approximately 1/5 (21%-23%) of the cancer types. Even more interesting is that blood microbiome appears more sensitive than SN, almost equally sensitive as PT, in demonstrating the heterogeneity associated with tumor.

TPLoN model parameters *a*, *b*, and *M*_0_ can be utilized to measure community (strictly, metacommunity) heterogeneity criticality threshold (HCT) (Ma 1991, 2005), where *M*_0_ = 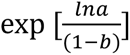, is the critical mean connectedness, at which the ratio of variance to mean (*V*/*M*) equals (&’() one, which implies that heterogeneity is random (possibly following Poisson distribution), similar to Eisler *et al*. (2008) endogenous behavior as determined by the system’s internal collective fluctuations. In other words, the external driving force (impact) such as environment influences are insignificant in driving the system (community) heterogeneity. When the mean connectedness (*M*) exceeds *M*_0_, the external driving force is significant in determining community heterogeneity. When (*M*<*M*_0_), the heterogeneity is even weaker than random distribution and is likely to follow uniform statistical distribution.

Compared with the previous heterogeneity analysis with TPLE, the tumor microbiome heterogeneity captured here with the TPLoN are the heterogeneity from species interactions, which should be more stable than those from species abundances. For this, the TPLoN scaling parameter (*b<2*) values are smaller than those of the TPLE parameter (*b*≥2) which represent the heterogeneity from both abundances and interactions. Furthermore, here the HCR (heterogeneity-cancer relationship) of species interactions captured with TPLoN is only 9%-12%, matching the lower interval of 10%-40% range of general HCR captured with TPLE in the previous sub-section.

## 5. Stochasticity analysis of the tumor microbiomes

We use Ning *et al*. (2019) NSR (normalized stochasticity ratio) framework to assess the stochasticity of community (metacommunity). A larger value of the NSR indicates higher stochasticity or lower selection level. Factors such as tumors may exert selection pressure on community assembly and may lower community stochasticity. Ning et al. (2019) NSR framework also includes a metric of community similarity, which is a kind of beta-diversity. The NSR similarity is an intermediate statistic in the NSR framework and is used to derive NSR: lower similarity implies higher beta-diversity. Both NSR and similarity range between 0 and 1, with NSR of 0 indicating pure deterministic community and 1 indicating pure stochastic community.

Table S24 exhibits the NSR values of tissue microbiome of each cancer types under different sequencing protocols and taxa (see Fig 7 for WGS protocol). Table S25 lists the results of Wilcoxon tests for the difference in NSR between PT and SN or between PT and B. Table S26 summarizes the Wilcoxon test results of the differences in NSR between different MSS sites, in terms of the percentages with significant differences. The percentages with significant differences in the NSR range from 50%-100%, mostly 70%-80% between normal tissues (B or SN) and PT. This suggests that the community (strictly speaking, metacommunity) level stochasticity is significantly different between MSS, especially between the normal tissues (SN or B) and PT, which should be expected.

**Fig 7.**
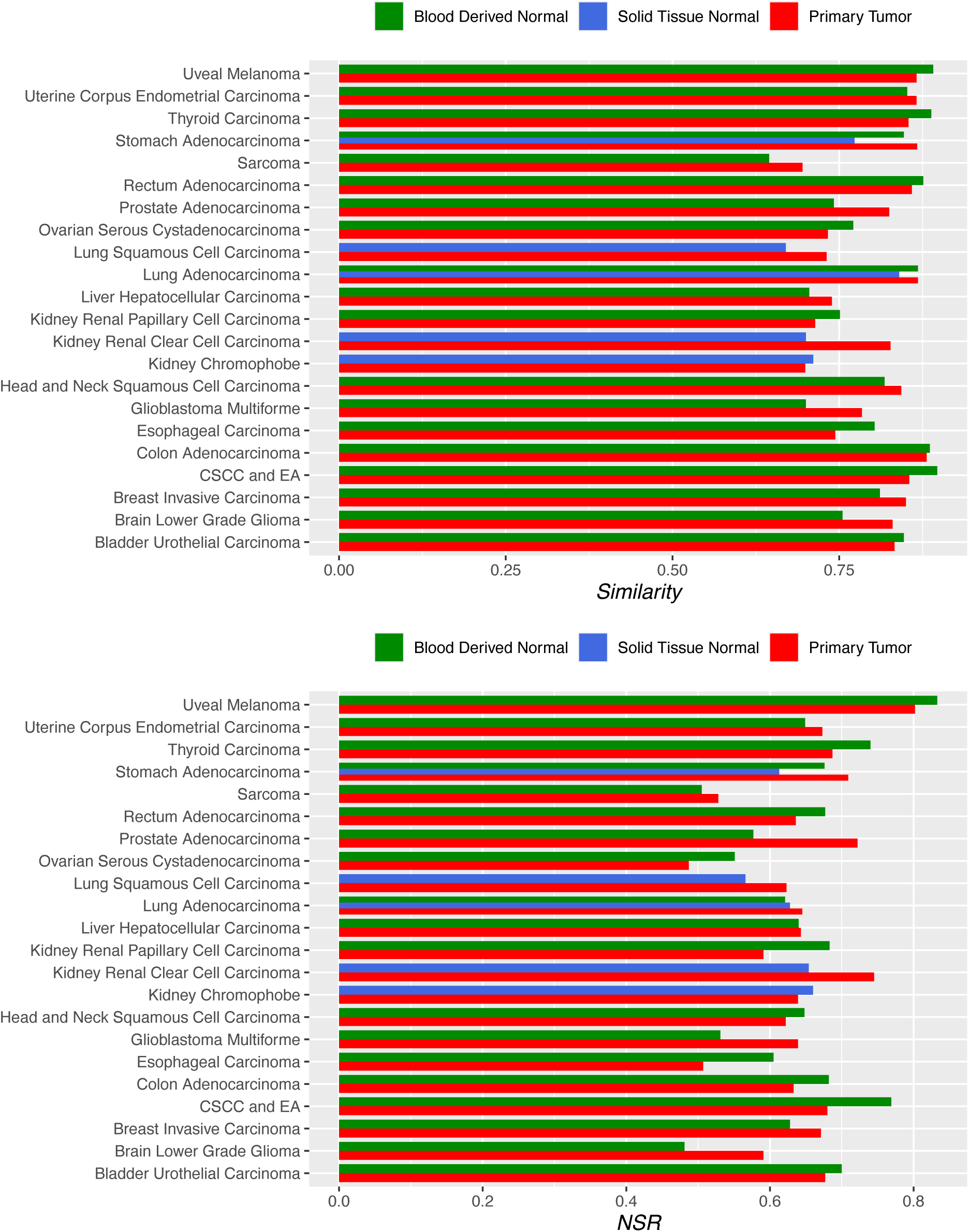
The comparisons of NSR (normalized stochasticity ratio) between PT (primary tumor), SN (solid tissue normal) and blood derived normal (B) for various cancer types with WGS datasets.

Finally, the range of NSR, approximately between 0.40 and 0.80 (NSR=[0.4, 0.8]), and especially exceeding 0.5 (NSR>0.5) in most comparisons (12 out of 16 or 75%) (Table S26) suggests that the community (metacommunity) structures and stability are mostly maintained by deterministic selections, of which tumor must be one of the major selection forces.

## 4. Conclusions and Discussion

The present study is aimed to investigate the tumor and pan-tumor microbiome diversity and heterogeneity of cancer tissue microbiomes with a big dataset distilled previously from TCGA database by Poore *et al*. (2020). The reanalysis of the microbiome data with a series of medical ecology approaches revealed the following findings:

(1) The tumor diversity-cancer relationship (DCR) is consistent with the 1/3 hypothesis of diversity-disease relationship (DDR) in human microbiome associated diseases (Ma *et al*. 2019). That is, in approximately 1/3 of cases, the microbiome diversity is significantly different between PT microbiomes and normal tissue microbiomes. Our study here revealed approximately 30%-40% differences between normal tissues (SN or B) and PT across 32 cancer types. As to the direction of DCR relationships, whether cancer enriches or deplete diversity, may depend on specific cancer types and progression stages, host physiology and genomes, lifestyles and diet habits, *etc*.

(2) The pan-tumor DCR varies greatly among cancer types, and the variations between cancer types range between 70%-80%, almost twice the percentage of the previous tumor-DCR.

(3) Tumor microbiome heterogeneity analysis with network diversity concept (metrics) of Ohlmann *et al*. (2019), which might be a misnomer given that its NDLA (network diversity in link abundances) and NDLP (network diversity in link probability) essentially measure the heterogeneity in a network setting, shows the tumor-HCR (heterogeneity-cancer relationship) of (0-100%) between normal (SN or B) and PT microbiomes. But in most cases, the tumor-HCR range (10%-36% with RNA-Seq data) is similar with previous tumor-DCR and is wider than the range of general HDR (heterogeneity-disease relationship) of microbiome-associated diseases (Ma 2020). The zero differences (0%) occurred mostly in the higher heterogeneity orders (q=1-3) of NDLP and NDLA with WGS datasets; and the bigger difference (40%-100%) occurred with *q*=0 in the WGS datasets, that is, the number of links in the network. The wider range of HCR than general HCR (Ma 2020) may has to do with the ‘direct’ nature of tumor microbiome: microbes are in direct contact with or even live inside tumor cells, while the previous HDR analyzed the relationship between gut microbiome heterogeneity and microbiome-associated diseases. This point is also applicable for the following pan-tumor heterogeneity comparisons, and in fact, also for the DCR.

(4) Tumor heterogeneity analysis with classic TPLE (Taylor’s power law extensions) revealed that, the tumor microbiome heterogeneity differences between PT and normal controls (B or NT) range between 10%-40% at community scale and 20%-50% at population scale. The percentage and range of heterogeneity differences are similar to the ranges of tumor microbiome diversity, but they measure different aspects of tissue microbiomes. The finding here is also consistent with the previous finding based on the network diversity with RNA-Seq data.

(5) The pan-tumor microbiome heterogeneity analysis with TPLE parameter (*b*) showed approximately 30%-70% differences between different cancer types, which is almost twice the differences of the previous tumor-level microbiome heterogeneity with TPLE parameter (b) (10%- 40%), which measures the microbiome heterogeneity between PT and normal (SN or B) microbiomes. That is, pan-tumor HCR is likely to be twice significant than tumor microbiome HCR. This is also similar with the comparison between pan-tumor diversity-cancer and tumor diversity-cancer relationships (i.e., pan-tumor DCR and tumor DCR). Furthermore, the pan-tumor microbiome heterogeneity scaling parameter (*b*) of PT is larger (higher end of the interval 70%) than that of B or SN (lower end of the interval 30%). This should be expected since PT should exert stronger selection pressure than normal tissues (SN or B) on its tissue microbiomes.

(6) Since classic TPL/TPLE measures heterogeneity by measuring the consequence of fluctuations in species abundances and interactions, rather than directly measuring species interactions, we extended TPL onto species cooccurrence networks (TPLoN) by fitting the classic TPL with the mean and variance of species connectedness in network setting (see Box 1). It was found that the TPLoN scaling parameter was only significantly different between PT and normal tissue (SN or B) in approximately 9%-12% of the comparisons across 32 cancer types. The percentage of TPLoN is on the lower end of the classic TPLE-*b* interval (10%-40%), which is somewhat puzzling! We postulate that the lower “effect size” captured with TPLoN is likely due to the reality that TPLoN directly captures the heterogeneity scaling of species interactions, which is likely to be smaller than those captured by classic TPL/TPLE that are built upon the fluctuations (variances) of both abundances and interactions. That is, interaction relationships are likely more stable (less heterogenous) than fluctuations of species abundances, not to mention the fluctuations of both. Yet, there is another alternative possibility that the proposed TPLoN might have introduced unnecessarily complexity, or “more is less!” We tend to believe that the first hypothesis is more plausible than the alternative conjecture.

(7) The NSR (normalized stochasticity ratio) analysis suggested that the percentages with significant differences in NSR range from 50%-100%, mostly 70%-80% between normal tissues (B or SN) and PT. That is, the community (strictly speaking, metacommunity) level stochasticity is significantly different between microbiome sites especially between the normal tissues and PT, which should be expected. Therefore, tumor growth should be a significant selection force in shaping the tissue microbiomes, leading to wide differences between PT and normal tissues. Since the range of NSR, approximately between 0.40 and 0.80, and especially exceeding 0.5 in most comparisons (12 out of 16 or 75%) further confirms that the community (metacommunity) is mostly regulated by deterministic selection, of which tumor must be one of the major selection factors.

(8) We postulate that either DCR or HCR is unlikely to be monotonic: whether tumors enrich or deplete diversity, raise or lower heterogeneity may depend on cancer types, cancer stage progressions, host physiology and genomics, lifestyle and diet habits, *etc*. One plausible interpretation is that selection forces such as tumors are largely deterministic, and they may either favor or disfavor certain species (or species groups) over others, which may either enrich or deplete species diversity, raise or lower heterogeneity. The findings in this study regarding tumor- and pan-tumor DCR (HCR) are consistent with the general DDR (diversity-disease relationship) and HDR (heterogeneity-disease relationship) of microbiome associated diseases (Ma *et al*. 2019, Ma 2020), and their differences are that in the case of cancer microbiomes, the range of differences in heterogeneity are wider than in diversity, opposite with the comparison between DDR and HDR.

(9) In tumor research, tumor heterogeneity is generally considered as a challenge from treatment perspective (Marusyk *et al* 2020 & Peer *et al*. 2021, Kashyap *et al* 2022). In other words, in most cases, taming tumor heterogeneity is likely favorable for managing cancers. The variable HCR/DCR suggests a different perspective in the case of tumor microbiome heterogeneity. In other words, heterogeneity can be a double sword for managing cancers. Therefore, variable or dynamic DCR/HCR patterns can provide both challenges and opportunities for managing the cancer-microbiome ecosystems towards healthy trajectory by measures such as using probiotics.

Finally, we discuss one observation: cancer research appears to be one of few fields that pay more attention on heterogeneity than on diversity, which is fully justified because of the importance of cellular interactions in carcinogenesis and cancer progression (*e.g.,* Kashyap *et al*. 2022). The findings of our study indeed suggest that the extent of microbiome heterogeneity differences is indeed wider than that of microbiome diversity differences at both tumor and pan-tumor levels, which suggests that heterogeneity is indeed more important than diversity in cancer microbiome research. In addition, much of existing studies on tumor heterogeneity are still focused on intra-tumor heterogeneity, but the importance of intertumor heterogeneity and pan-tumor heterogeneity should not be ignored, which is particularly true in the case of tumor microbiomes. Intuitively, unlike tumor cells, microbiomes are virtually everywhere within our bodies and on our skins, and they are our residents from our births. Limiting microbiome research within a single tumor, or the so-termed intra-tumor microbiome heterogeneity, a term we did not adopt in this study, is not unlike focusing on islands in ocean or trees in forest. Using the terminology in tumor heterogeneity research (Marusyk *et al*. 2020), none of the previous contents we presented could be classified into the intra-tumor heterogeneity paradigm. We hope the approaches we demonstrated in this study will also be applied to investigate general inter-tumor heterogeneity or pan-tumor heterogeneity beyond microbiomes, and indeed most of the approaches should be equally applicable for investigating cellular, genomic and epigenetic heterogeneities in tumor heterogeneity research.

## Data Availability

https://doi.org/10.1038/s41586-020-2095-1

## Online Supplementary Informa2on

Table S1-S26 (in Separate MS-Excel File)

## Acknowledgements

This study received funding from the following sources: A NaFonal Natural Science FoundaFon (NSFC) Grant (No. 31970116, 72274192). I appreciate the computaFonal support from Dr. Lianwei Li of Chinese Academy of Sciences.

## Availability of datasets

Poore, GD, E Kopylova, Q Zhu, … R Knight (2020). Microbiome analyses of blood and Fssues suggest cancer diagnosFc approach. *Nature* 579, 567–574. https://doi.org/10.1038/s41586-020-2095-1

## Ethic Approval

N/A since the study does not involve any wet-lab experiments or survey on human or animal subjects, and all analyzed datasets are already available in public domain, as menFoned above.

## References

Chao, A., Chiu, CH. & Jost, L. (2014) Unifying species diversity, phylogenetic diversity, functional diversity and related similarity and differentiation measures through Hill numbers. Annual Reviews of Ecology, Evolution, and Systematics, 45, 297–324.

Eisler Z, Bartos I, Kertész J (2008) Fluctuation scaling in complex systems: Taylor’s law and beyond. Advances in Physics 57:89–142.

Gough, A., Stern, A. M., Maier, J., Lezon, T., Shun, T. Y., Chennubhotla, C., … & Taylor, D. L. (2017). Biologically relevant heterogeneity: metrics and practical insights. SLAS Discovery: Advancing Life Sciences R&D, 22(3), 213–237.

Hanahan D. (2022) Hallmarks of Cancer: New Dimensions. Cancer Discov. Vol.12(1):31–46. doi: 10.1158/2159-8290.CD-21-1059.

HMP Consortium. (2012) Structure, function and diversity of the healthy human microbiome. Nature 486:207–214.

Hill, M. O. (1973) Diversity and evenness: a unifying notation and its consequences. Ecology, 54:427–342.

Integrative HMP (iHMP) Research Network Consortium (2019) The Integrative Human Microbiome Project. Nature 569:641–648.

Lythgoe MP, Mullish BH, Frampton AE, Krell J. (2022) Polymorphic microbes: a new emerging hallmark of cancer. Trends Microbiol. Vol. 30(12):1131–1134. doi: 10.1016/j.tim.2022.08.004.

Kashyap, A et al. (2022) Quantification of tumor heterogeneity: from data acquisition to metric generation. Trends in Biotechnology, June 2022, Vol. 40(6):648–676

Ma ZS (2015) Power law analysis of the human microbiome. Molecular Ecology, 24(21):5428–5445.

Ma, Z. S and A. M. Ellison. (2018) A unified concept of dominance applicable at both community and species scale. Ecosphere, 10.1002/ecs2.2477.

Ma ZS, Ellison AM (2019) Dominance network analysis provides a new framework for studying the diversity-stability relationship. Ecological Monographs. 89(2), DOI: 10.1002/ecm.1358.

Ma ZS, Li LW, Gotelli NJ (2019) Diversity-disease relationships and shared species analyses for human microbiome-associated diseases. The ISME Journal, 13: 1911–1919.

Ma, ZS (2020). Heterogeneity-disease relationship in the human microbiome associated diseases. FEMS Microbiology Ecology, Vol. 96, fiaa093. doi:10.1093/femsec/fiaa093.

Ma ZS & Taylor RAJ (2020) Human reproductive system microbiomes exhibited significantly different heterogeneity scaling with gut microbiome, but the intra-system scaling is invariant. Oikos, 129 (6): 903–911.

Ma ZS (2021) Cross-scale analyses of animal and human gut microbiome assemblies from metacommunity to global landscape. mSystems 6: e00633–21. 10.1128/mSystems.00633-21.

Marusyk, A., Janiszewska, M., & Polyak, K. (2020). Intratumor heterogeneity: the rosetta stone of therapy resistance. Cancer Cell, 37(4), 471–484.

Ning, D., Deng, Y., Tiedje, J.M., Zhou, J. (2019). A general framework for quantitatively assessing ecological stochasticity. PNAS, 116 (34): 16892–16898.

Ohlmann M, Miele V, Dray S, et al. (2019) Diversity indices for ecological networks: a unifying framework using Hill numbers. Ecology Letters, 22(4): 737−747.

Peer D et al. (2021) Tumor heterogeneity. Cancer Cell Vol. 39, 1015–1017

Poore, GD, E. Kopylova, Q Zhu, … R Knight (2020). Microbiome analyses of blood and tissues suggest cancer diagnostic approach. Vol. 579, 567–574. 10.1038/s41586-020-2095-1

Sepich-Poore, GD, L Zitvogel, R Straussman, J Hasty, JA Wargo, and R Knight (2021) The microbiome and human cancer. Science, vol. 371, eabc4552 (2021).

Shavit, Ayelet & Ellison, Aaron M. (2021). Diverse Populations are Conflated with Heterogeneous Collectives. Journal of Philosophy, Vol. 118 (10):525–548.

Taylor LR (1961) Aggregation, variance and the mean. Nature 189:732–735.

Taylor LR (1984) Assessing and interpreting the spatial distributions of insect populations. Annual Review of Entomology 29:321–357.

